# Cognitive Enhancement in Bipolar Disorder: A Double-Blind, Randomized Controlled Trial Utilizing a Novel DTI-Guided Multimodal Neuro-stimulation Protocol

**DOI:** 10.1101/2024.07.25.24311037

**Authors:** Minmin Wang, Xiaomei Zhang, Hetong Zhou, Qianfeng Chen, Qiqi Tong, Qiai Han, Xudong Zhao, Dandan Wang, Jianbo Lai, Hongjian He, Shaomin Zhang, Shaohua Hu

**Author notes:** These authors contributed equally (Minmin Wang, Xiaomei Zhang, Hetong Zhou). **Corresponding authors: Shaohua Hu**, Department of Psychiatry, the First Affiliated Hospital, Zhejiang University School of Medicine, Hangzhou, China.

## Abstract

**Background:** Traditional neuromodulation strategies for enhancing cognitive abilities in bipolar disorder (BD) patients have shown promise, yet there remains a need for novel intervention modalities to improve therapeutic outcomes.

**Methods:** This study introduces a novel multi-modal neuro-stimulaton (MNS) protocol using individualized DTI data to identify fiber tracts between the DLPFC and dACC. The highest structural connectivity point is selected as the individualized stimulation target, which is targeted using a combination of optimized tACS and robot-assisted navigated rTMS. A double-blind randomized controlled trial (Trial registration number: NCT05964777) was conducted to investigate the clinical efficacy of this innovative neuromodulation approach on cognitive abilities in BD patients. One hundred BD patients were randomly assigned to four groups: Group A (Active tACS-Active rTMS (MNS Protocol)), Group B (Sham tACS-Active rTMS), Group C (Active tACS-Sham rTMS), and Group D (Sham tACS-Sham rTMS). Participants underwent 15 sessions over three weeks. Cognitive assessments (THINC integrated tool) were conducted at baseline (Week 0), post-treatment (Week 3), and follow-up (Week 8).

**Results:** Sixty-six participants completed all 15 sessions. Group A (MNS Protocol) showed superior improvements in Spotter CRT, TMT, and DSST scores compared to other groups at Week 3, with sustained cognitive enhancement in Spotter CRT at Week 8 (*P* < 0.01). Only Group A exhibited significant activation in the left frontal region post-MNS intervention. The novel MNS protocol was well tolerated, with no significant side effects observed.

**Conclusions:** DTI-guided multimodal neuro-stimulation mode significantly improves cognitive impairments and is safe for BD patients.

**Highlights:**

1. Using DTI-derived neural fiber density to determine the target sites for tACS and rTMS in patients with bipolar disorder.
2. Combining sequential tACS and rTMS to significantly improve cognitive function in the bipolar disorder patients.
3. Utilizing individually optimized tACS and robot-assisted navigated rTMS to achieve high-precision transcranial stimulation.

## 1. Introduction

Bipolar disorder (BD) is a complex and debilitating psychiatric disease characterized by recurrent episodes of mania and depression^1^. Patients with BD may suffer from widespread cognitive impairments, including deficits in attention, learning, memory, and executive functions, leading to significant occupational and social dysfunction^2^. Current treatments for cognitive impairment in BD, including pharmacological, psychological, and behavioral therapies, are constrained by heterogeneous treatment outcomes, slow symptom relief, and associated risks and side effects^3,4^. These limitations highlight the need for innovative therapeutic approaches to improve the cognitive functions^5,6^.

Non-invasive neuromodulation, such as transcranial electrical stimulation (tES) and repetitive transcranial magnetic stimulation (rTMS), has emerged as promising methods for cognitive enhancement in patients^7,8^. Transcranial alternating current stimulation (tACS), in particular, has shown great potential due to its unique rhythmic modulation capabilities in the treatment of neuropsychiatric disorders^9,10^. The tACS may resynchronize intrinsic brain rhythms, thereby modulating cognitive processes^11^. Studies have demonstrated that 40 Hz tACS applied to the dorsolateral prefrontal cortex (DLPFC) can improve cognitive function in patients with psychiatric disorders and has also shown beneficial effects in cognitive enhancement in healthy subjects^12–15^. Additionally, 40 Hz tACS has exhibited promising applications^16^. Despite these advances, conventional tES primarily uses a two-electrode stimulation mode, which has limitations in terms of facality. Although a 4×1 ring electrode array was proposed to enhance stimulation facality, individual variations in brain tissue morphology significantly influence the distribution of electrical currents within the brain^17^. Recent studies have proposed using individualized simulation models of tES and electrode optimization methods to enhance the electric field (EF) intensity in target brain regions, minimizing current spread to non-target areas^18,19^. Consequently, individualized 40 Hz tACS holds promise for significantly improving cognitive function in BD patients during remission.

Moreover, high-frequency rTMS of the DLPFC has been reported to positively impact memory and executive function in BD patients^20,21^. However, the clinical efficacy of rTMS for cognitive impairments in BD was inconsistent^22–24^, possibly due to variability in target selection and accuracy of localization^25,26^. Traditional rTMS often failed to consider structural and functional differences between individuals, resulting in suboptimal stimulation outcomes and sometimes missing the abnormal target regions^27^. Thus, achieving precise stimulation is crucial for enhancing the efficacy of rTMS^28^. Stanford University introduced the SNT mode, marking the first clinical application of individualized functional connectivity-guided TMS, targeting the strongest functional connectivity points between the subgenual anterior cingulate cortex (sgACC) and DLPFC^29,30^. The approach improved connectivity between deeper brain regions and the DLPFC, emphasizing the critical role of neuro-navigation in achieving accurate target localization and demonstrating the potential of personalized rTMS intervention modes^31^. Transitioning from these single-mode brain stimulation techniques, combining tES and rTMS has been shown to potentially enhance cortical activity regulation and cognitive function^32–34^, although limited studies have explored the synergistic effects of combined electromagnetic stimulation on clinical outcomes^35–37^. The combined effects of tACS and rTMS on BD patients remain an area requiring further investigation.

Additionally, target selection presents another significant challenge in neuromodulation for BD. While traditional approaches focus on structural targets, current methods are exploring the use of individual functional connectivity to identify specific targets^29,38^. However, results have shown that functional targets can be influenced by the processing of resting-state MRI data, leading to variability in target identification and raising questions about the stability of such methods. Furthermore, the effectiveness of interventions based on functional targets remains to be fully explored^39,40^. On the other hand. structural imaging studies have revealed a disrupted white matter microstructure as a potential pivotal clues in BD^41,42^. Several studies have indicated decreased white matter integrity, reduced fractional anisotropy (FA), and lower levels of axonal myelination, particularly in the fronto-subcortical circuits^43,44^. Moreover, certain frontal-cingulate white matter connections have also been identified as stimulation targets of deep brain stimulation for major depressive disorder (MDD), demonstrating their therapeutic potential^45,46^. These findings suggest that using white matter structural connectivity to determine stimulation targets may be an effective approach. Structural pathways are likely to facilitate the transmission of physical intervention effects, suggesting that neuromodulation based on structural connectivity could yield better outcomes compared to functional connectivity-based approaches.

This study aims to introduce a novel multimodal neuro-stimulation protocol (MNS Protocol) that utilizes individualized DTI data for target localization, and combines individually optimized tACS followed by robot-assisted navigated rTMS for multimodal intervention. We conduct a double-blind randomized controlled trial to validate the effects of the novel neuro-stimulation mode on the cognitive impairment of BD patients. The findings of this research are expected to advance the field of neuromodulation therapies for cognitive impairment in BD.

## 2. Methods and Materials

### 2.1 Novel Multimodal Neuro-stimulation Protocol (MNS Protocol)

As shown in Figure 1, the novel MNS Protocol is a sequential stimulation approach that combines personalized tACS and robot-assisted navigated rTMS. The personalized tACS intervention is administered at 40Hz for a duration of 30 minutes. Immediately following the tACS, neuro-navigated rTMS is performed. This protocol leverages individualized DTI data to precisely identify fiber tracts between the DLPFC and dACC, selecting the point of highest structural connectivity as the stimulation target. By optimizing stimulation parameters for both tACS and rTMS based on individual head model, the MNS Protocol aims to enhance cognitive function in BD patients by utilizing advanced neuroimaging and targeted stimulation techniques. The specific method is described as follows:

**Figure 1.**
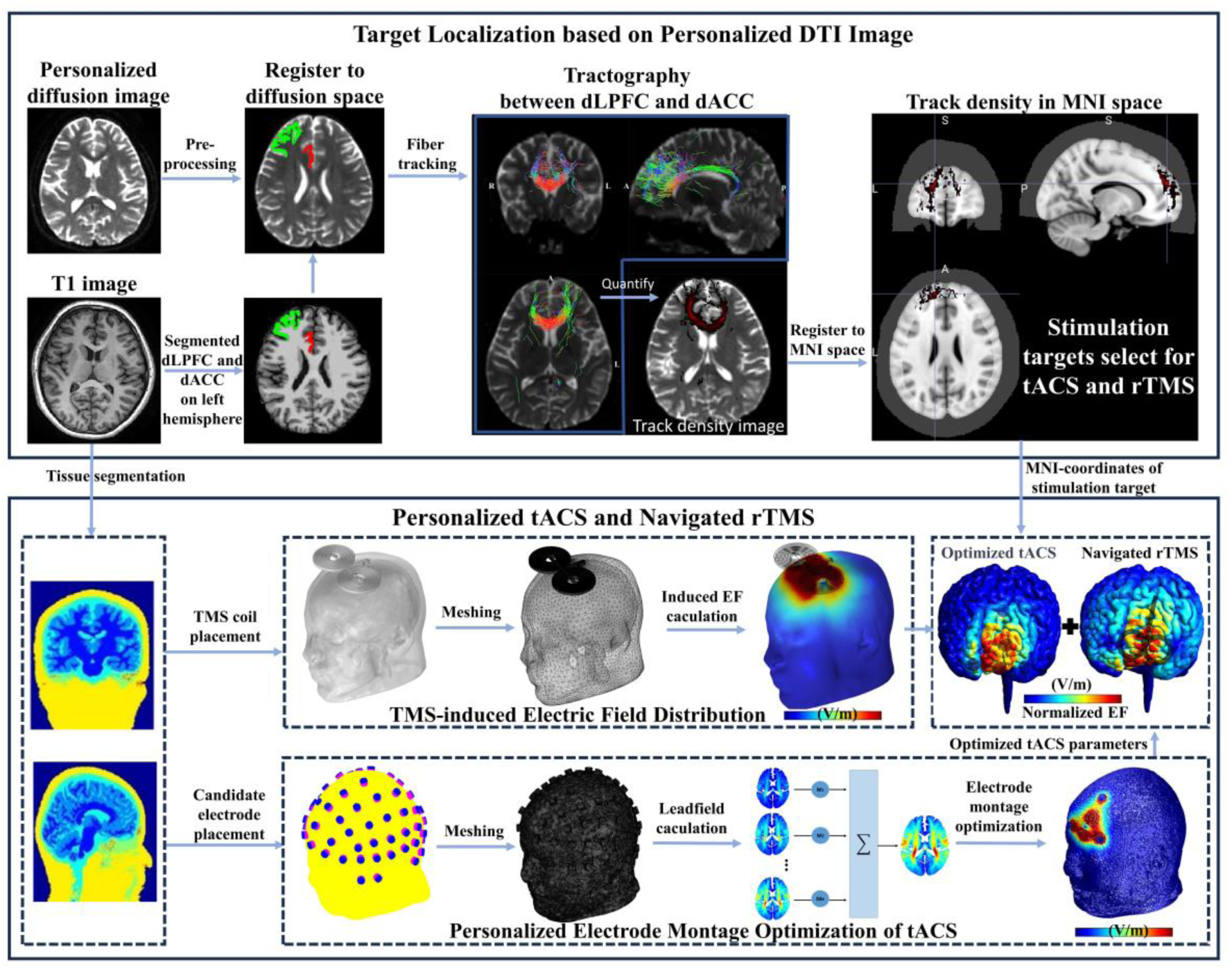
Schematic diagram of multimodal neuro-stimulation protocol. Individual DTI data are analyzed to determine the neural fiber tract density between the DLPFC and dACC. The point with the highest connectivity density is identified as the stimulation target. Using individual T1-weighted structural images, personalized induced EF models for tACS and navigated rTMS are constructed. The tACS parameters are then optimized ensuring precise and effective stimulation in the stimulation target,, followed by the application of navigated rTMS.

#### 2.1.1 Target Localization Based on Individualized DTI Image

For the individualized target localization based on DTI imaging, as shown in Figure 1, T1-weighted structural images and DTI data were acquired from patients and subsequently preprocessed. The detailed scanning parameters can be found in section 2.5. The processing of T1 images used Freesurfer software for automated region-of-interest segmentation, including image grayscale normalization, gray and white matter segmentation, and fine cortex segmentation using the Desikan-Killiany template. The DTI images were processed using MRtrix3 and FSL software, including image noise reduction, Gibbs artifact removal, eddy current correction, etc. Secondly, after calculating the response function and CSD fiber directional distribution function, a fiber tracking algorithm based on the second-order FOD was used to reconstruct one million fibers in the whole brain. Subsequently, the two pre-segmented regions of interest DLPFC and dorsal anterior cingulate cortex (dACC) in the T1 structural image were extracted, registered into the DTI space, the fiber bundles passing through the two regions at the same time were screened, and the fiber density in each voxel in the fiber path was calculated as a measure of the structure. An indicator of the strength of the connection. Finally, the voxel position corresponding to the highest point of structural connection strength in the individual DTI space was converted to the MNI152 (Montreal Neurological Institute) standard T1 template space after nonlinear registration as the stimulation target.

#### 2.1.2 Treatment Protocol of Personalized tACS

After determining the individualized stimulation target, we proceeded to optimize the treatment parameters of tACS for each patient as depicted in Figure 1. We optimize the electric field intensity and focus of the target point. While maximizing the electric field intensity in the target area, it minimizes the electric field intensity in non-target areas and improves the focus of the electric field. In addition, some current constraints are set for safety reasons, including the size limit of the total electrode current and the size limit of a single electrode current. The specific steps are as follows:

First, the SIMNIBS tool software was applied for constructing individualized head models. The selected stimulation electrode employed in this study is a 10 mm diameter disc electrode. The electrical conductivity characteristics of each corresponding tissue and material set to: 0.465 S/m (scalp), 0.01 S/m (skull), 1.65 S/m (cerebrospinal fluid), 0.276 S/m (gray matter), 0.126 S/m (white matter), 2.5e-14 S/m (air cavity), 5.9e7 S/m (electrode), and 0.3 S/m (conductive paste).

The proposed electrode optimization approach for tES involves the following sequential steps. Primarily, the initial step involves the calculation of the leadfield matrix, denoted as *A*:

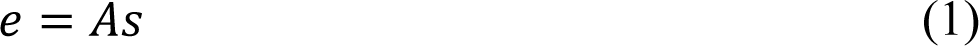

Where:

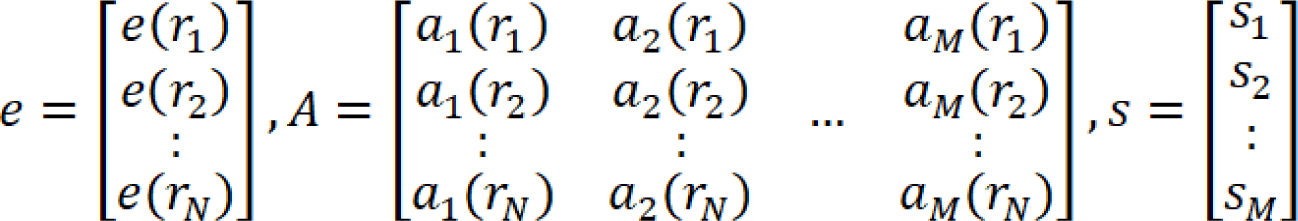

*r_N_* represents the node position on the head model, *N* denotes the number of nodes in the head model, and *M* signifies the total number of candidate electrodes.

To enhance computational efficiency, the EF intensity of the minimized non-target region is quantified as the EF intensity on the voxel point situated at a distance of 1/4* the maximum distance from the target point. The overarching objective of electrode optimization is expressed through the optimization function:

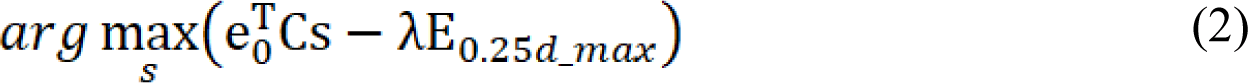

where *s* denotes the magnitude of the stimulation current*, e ^T^C* signifies the distribution of the required EF intensity at the target, and E_0.25d_max_ denotes the EF intensity at a node located at 1/4* the maximum distance from the target point. The weight parameter *λ* is used to balance the two objectives of focality and EF intensity.

Constraints condition are integral to the optimization method. Firstly, the total input current of the electrode must equal the output current, ensuring a balanced current sum.

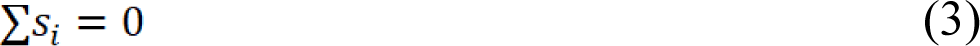

Additionally, the sum of the absolute values of the current is not larger than *I_total_*.

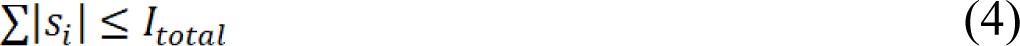

and the absolute value of the current of a single electrode is not greater than *I_max_*.

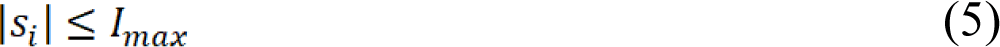

for clinical applicability, constraints limiting the number of electrodes (denoted as 𝑛) are introduced:.

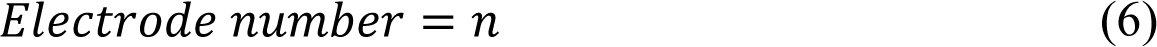

and the absolute value of the current of a single electrode is not greater than Imax.

In this study, we set the *I_total_* at 4 mA, with a maximum current *I_max_* of 2 mA per electrode. The number of electrodes 𝑛 was selected to be either 4 or 5.

#### 2.1.3 Treatment Protocol of Navigated rTMS

In this study, we combined the acquired individual structural MRI images to construct personalized head models using SIMNIBS software. Our head models encompassed essential components such as the scalp, skull, cerebrospinal fluid (CSF), gray matter (GM), white matter (WM), and other brain tissues. The figure-of-eight coil, featuring an inner diameter of 20 mm and an outer diameter of 70 mm, was integrated into the simulation model. The coil was positioned directly above the C3 region and simulated as current sources characterized by a frequency of 3 kHz and a current amplitude of 5 kA. We conducted simulations to analyze the electric fields generated within the brain by TMS at different coil angles. The optimal coil placement angle was selected to ensure that the electric field at the target site was sufficiently large for each individual.

A neuro-navigation system was employed for rTMS. Firstly, the individualized head model was co-registered with a standard brain using AC-PC alignment. The MRI images were converted into three-dimensional skin and brain models to replicate the head characteristics of patient. Using a visual localization system, a head positioning mask, and appropriate computer software, the treatment site was identified on the patient’s scalp. Subsequently, the treatment was administered by aligning the coil at the specified angle with the determined treatment site.

#### 2.1.4 Sequential Personalized tACS and Navigated rTMS

The MNS protocol proposed in this study represents a departure from traditional unimodal electrical or magnetic stimulation techniques. This protocol employs a dual-modal approach, combining personalized tACS with navigated rTMS. The personalized tACS intervention is administered at 40Hz for a duration of 30 minutes. Immediately following the tACS, neuro-navigated rTMS is performed. Each daily rTMS treatment consisted of 60 five-second 10-Hz trains delivered at 100% of the resting motor threshold (RMT) with inter-train intervals of 15 s (i.e., 3000 pulses per session, 45,000 pulses overall). For the sham treatment, the coil was directed to the same target with the simulation of scalp sensations with the production of the same sounds, without the application of the magnetic field. Each participant received 15 days of tACS-rTMS treatment. This sequential application is designed to maximize the synergistic effects of electromagnetic stimulation on cognitive function.

### 2.2 Trial Design

As illustrated in Figure 2A, to validate the efficacy of the proposed MNS protocol, a randomized controlled double-blind clinical trial was conducted. The stringent experimental design ensures that both participants and investigators remain blinded to the treatment allocation, thereby minimizing potential biases and enhancing the validity and reliability of the findings.

**Figure 2.**
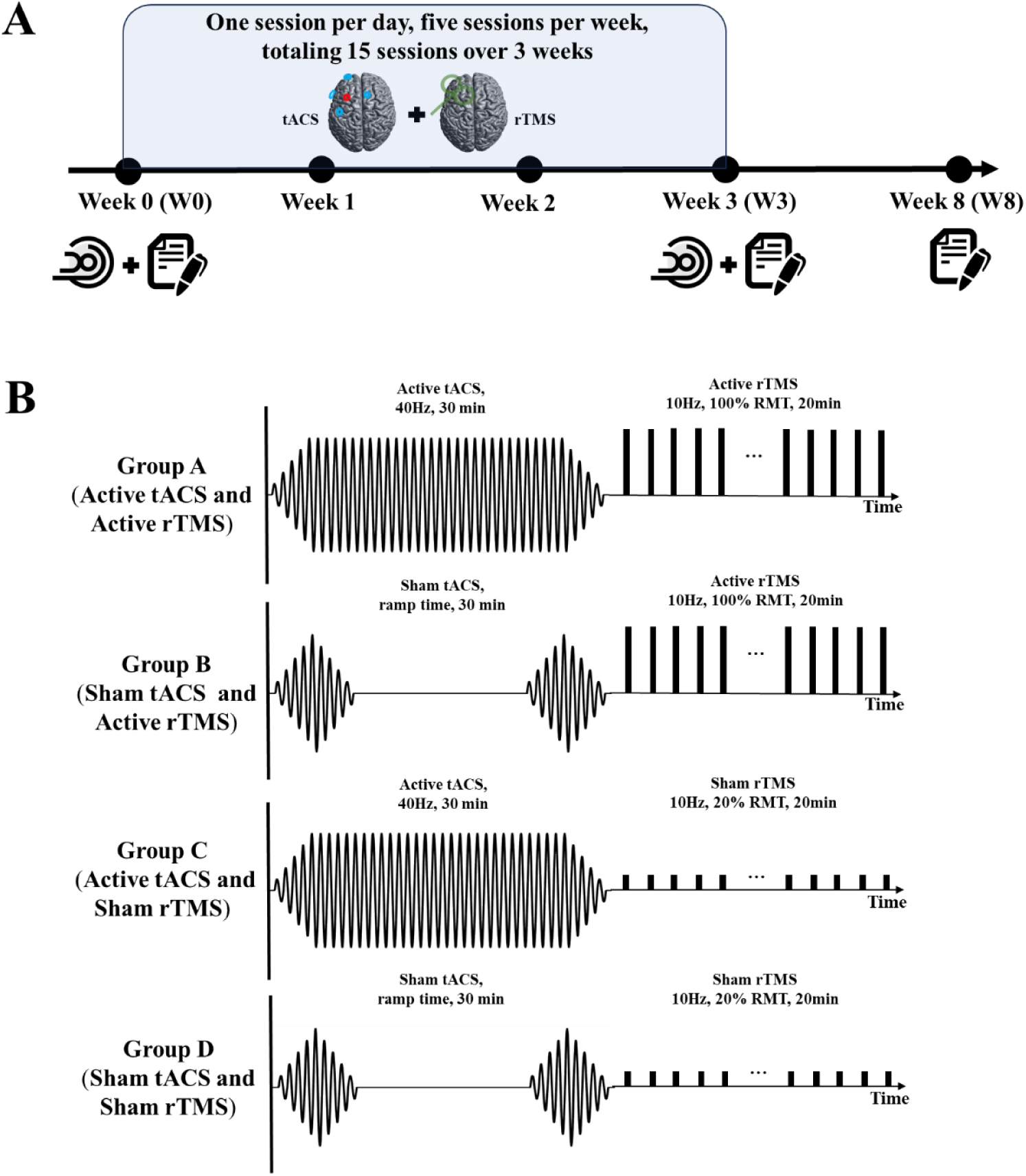
Clinical trial design and intervention paradigm. A. Participants underwent a three-week multimodal intervention combining transcranial electrical stimulation and transcranial magnetic stimulation. Each week consisted of daily sessions, totaling 15 sessions. Cognitive assessments and imaging examinations were conducted before and after the intervention, with a cognitive follow-up assessment in the fifth week post-intervention. B. Group A received 30 minutes of Active tACS followed by 20 minutes of Active rTMS. Group B received 30 minutes of Sham tACS followed by 20 minutes of Active rTMS. Group C received 30 minutes of Active tACS and Sham rTMS. Group D received 30 minutes of Sham tACS and Sham rTMS. Each participant with recorded scores at three time points, W0: Week 0, W3: Week 3, W8: Week 8.

Participants were randomly assigned to any one of the four groups: A, B, C, or D. In Group A (Active tACS and Active rTMS (MNS Protocol)), tACS is the active stimulation, and rTMS is the active stimulation group. In Group B (Sham tACS and Active rTMS), tACS is the sham stimulation, and rTMS is the active stimulation group. In Group C (Active tACS and Sham rTMS), tACS is the active stimulation, and rTMS is the sham stimulation group. In Group D (Sham tACS and Sham rTMS), both tACS and rTMS are sham stimulation. The personalized localization was conducted for the four groups of patients, All the participants were received a consecutive three-week treamtment including five times per week.

As illustrated in Figure 2B, the active rTMS group received rTMS treatment at 100% RMT, while the sham stimulation group received stimulation at 20% RMT. The active tACS group underwent 30 minutes of alternating current stimulation, whereas the sham tACS group had current passing only during the initial and final 30 seconds of the stimulation. Cognitive and imaging assessments were conducted before (Week 0, W0) and after (Week 3, W3) the treatment. Psychological scale evaluations were performed during the treatment, and adverse reactions were documented. The clinical cognitive symptoms and treatment safety were evaluated, with follow-up cognitive evaluations conducted at Week 8 (W8).

### 2.3 Participants

This study recruited patients with stable-phase BD and cognitive impairment from the outpatient Psychiatric Department of the First Affiliated Hospital, Zhejiang University School of Medicine, from July 2023 to February 2024. The research was approved by the Clinical Research Ethics Committee of the First Affiliated Hospital, Zhejiang University School of Medicine (Approval number: IIT20230058C-R1). The study has been registered in the Clinical Trials Registry (http://www.clinicaltrials.gov) with the registration number NCT05964777. Before participation, all participants provided informed consent, ensuring their understanding of the objectives, procedures, and potential risks of this study. Participant confidentiality and well-being was upheld throughout the study.

The inclusion criteria included, (i) Aged 14-45, regardless of gender; (ii) Diagnosis of BD in accordance with the Mini-International Neuropsychiatric Interview (M.I.N.I.); (iii) Stable medication treatment for at least three months; (iv) Clinical remission for at least 3 months before entering the randomization phase, with a Young Mania Rating Scale (YMRS) score ≤6 and Hamilton Depression Rating Scale (HAMD-17) score ≤7; (v) Cognitive impairment indicated by a Perceived Deficits Questionnaire (PDQ) score ≥17; (vi) Right-handed; (vii) Education level ≥9 years.

The exclusion criteria included: (i) Comorbidities of any other mental disorder in the Diagnostic and Statistical Manual of Mental Disorders, Fifth Edition (DSM-5); (ii) Any contraindications for MRI or the discovery of structural abnormalities in brain MRI; (iii) Pregnant, lactating women, breastfeeding or those planning to become pregnant; (iv) Patients who have undergone ECT, rTMS, or tACS treatment in the last 3 months; (v) History of serious neurologic illnesses, such as epilepsy or traumatic brain injury; (vi) Significant and unstable medical conditions, including diabetes or cardiovascular, hematological, endocrine, liver, or kidney disease; (vii) History of substance abuse and dependence, (viii) Achromatopsia, hypochromatopsia, or dysaudia; (ix) Medication with antidepressant or anticholinergic agents.

### 2.4 Imaging

Imaging was conducted using a 3.0 Tesla scanner (GE SIGNA) with a standard whole-head coil at the First Affiliated Hospital, Zhejiang University School of Medicine. Participants were instructed to lie on the scanner bed with their eyes closed. Foam cushions were placed on either side of the head to minimize head motion, and ear plugs were provided to reduce noise interference. High-resolution 3D anatomical images were acquired using a T1-weighted magnetization prepared rapid acquisition gradient-echo (MPRAGE) sequence. The scanning parameters were as follows: 3D T1 structural image: TR/TE = 7.1/2.9 ms; 146 slices, slice thickness 1 mm; flip angle = 8°; matrix size = 256×256; field of view = 260×260 mm². For the DTI diffusion image, a planar echo sequence was used with the following parameters: TR/TE = 11000/89.4 ms; matrix size = 256×256; field of view = 240×240 mm²; number of slices = 44; slice thickness = 3 mm. The b value for DTI scanning was 1000 s/mm², and 25 diffusion directions were included. During scanning, participants were instructed to keep their eyes closed. To minimize head motion, foam cushions were strategically placed on either side of the head, and earplugs were utilized to mitigate noise.

### 2.5 Imaging Analysis

#### 2.5.1 fMRI Analysis

The fMRI data preprocessing was conducted using Data Processing Assistant for Resting-State fMRI (DPARSF) Advanced Edition V5.4 in MATLAB 2020. The key preprocessing steps included segmenting the MRI data into 10 periods, discarding the first 10 noisy time points, and synchronizing slice timing. Participants with head movement exceeding 3 mm or 3°were excluded from the analysis. Functional images were normalized to the MNI space using T1-based segmentation and registration. Spatial smoothing was applied using an 8mm×8mm×8mm Gaussian kernel. Linear drift was removed through linear regression to reduce the effects of thermal noise. Band-pass filtering (0.01-0.1Hz) was employed to eliminate physiological noise. The brain was parcellated into regions based on the AAL116 template, treating each region as a network node. Pearson correlation coefficients of the time series were computed to construct brain network matrices, which were Fisher Z-transformed for subsequent statistical analysis. Graph theory analysis was conducted using GRETNA software, calculating global and nodal metrics of the brain network matrices for both the baseline and post-treatment groups over a sparsity range of 0.01<S<0.35 with a step size of 0.01. Global metrics included the small-world index, normalized characteristic path length, characteristic path length, clustering coefficient, global efficiency, local efficiency, and network efficiency. Nodal metrics comprised nodal efficiency, nodal local efficiency, nodal clustering coefficient, nodal shortest path, betweenness centrality, and degree centrality.

#### 2.5.2 DTI Analysis

The DTI data were processed using FSL software (FMRIB Software Library, Oxford, UK), including motion and eddy current correction, brain extraction, tensor model fitting, and normalization to the MNI space using Tract-Based Spatial Statistics (TBSS). First, motion and eddy current corrections were applied to each participant’s diffusion-weighted imaging (DWI) data, and the diffusion gradient directions were rotated accordingly using affine registration. Subsequently, the diffusion tensor model was fitted using a weighted least squares approach in each participant’s native space. Spatial normalization involved mapping each participant’s FA image to a standard space (2mm isotropic MNI152 template) using non-linear registration. The FA reference and subsequent region of interest (ROI) definitions were based on the ICBM-DTI-81 white-matter atlas from Johns Hopkins University. Finally, scalar metrics from 40 ROIs were extracted from each participant’s projected skeleton.

### 2.6 Clinical Assessment

The Depression Perceptual Deficit Questionnaire (PDQ) was utilized to screen for cognitive impairment in participants at baseline. The severity of symptoms was evaluated using the 17-item Hamilton Depression Rating Scale (HDRS-17) and Young Mania Rating Scale (YMRS) at different time points. Assessors underwent training for consistency prior to assessments, and the adverse events was recorded.

The THINC integrated tool (THINC-it) is a self-administered iPad-based assessment tool that has been demonstrated to be reliable and valid for evaluating cognitive symptoms in BD patients. THINC-it consists of Spotter, SymbolCheck, Codebreaker, Trails, and 5 items from the Depression Perception Deficit Questionnaire (PDQ-5-D), corresponding to 5 traditional neuropsychological tests: the recognition task (CRT), Stroop task (1 back), the Digit Symbol Substitution Test (DSST), Digit Matching Line (TMT), and PDQ. These tests assess attention, processing speed, working memory, and cognitive and executive functions. Cognitive function in all participants was assessed using THINC-it at Week 0 (W0), Week 3 (W3), and Week 8 (W8) for each participant. Any adverse events encountered were meticulously documented.

The primary outcome measure was the change in scores on the five THINC-it tests between Week 0, Week 3 and Week 8. Secondary outcomes included changes in functional connectivity (FC) and structural connectivity between the DLPFC-dACC at the end of the 3rd week.

### 2.7 Clinical Outcome Analysis

The collected scores from the assessment tools were subjected to statistical analysis using repeated measures analysis of covariance (ANOVA) and paired t-tests. Repeated measures ANOVA was employed to assess within-group and between-group differences across the three-time points (W0, W3, and W8). Paired t-tests were used to investigate specific changes between baseline and post-treatment or follow-up assessments.

This study employed MATLAB 2020b, SPM12, and DPABI for statistical analysis. SPM12 was used to conduct paired t-tests to examine variations in fALFF, ReHo, and FC between groups. The DPABI software, in conjunction with Gaussian Random Field (GRF) theory, was utilized to correct for multiple comparisons in the paired t-test results. This correction included a voxel threshold of *P*<0.01 and a clump threshold of *P*<0.05.

## 3. Results

### 3.1 Clinicodemographic Characteristics of Patient

In this study, a total of 116 participants were initially recruited as presented in Figure 3. Sixteen were excluded, resulting in 100 BD patients who were ultimately enrolled and randomly assigned into four treatment groups in equal numbers: Group A (Active tACS-Active rTMS, n=25), Group B (Sham tACS-Active rTMS, n=25), Group C (Active tACS-Sham rTMS, n=25), and Group D (Sham tACS-Sham rTMS, n=25). Among these 100 participants, 66 completed all 15 planned MNS stimulation sessions and the assessments at Week 0 and Week 3 (Group A: 18 participants, Group B: 17 participants, Group C: 18 participants, Group D: 13 participants). Due to various clinical factors, only 37 participants completed the follow-up assessment at Week 8 (Group A: 13 participants, Group B: 10 participants, Group C: 7 participants, Group D: 7 participants). The demographic characteristics of these participants, including age, gender, and baseline symptom severity, showed no significant statistical differences across the groups (see **Table 1**).

**Figure 3.**
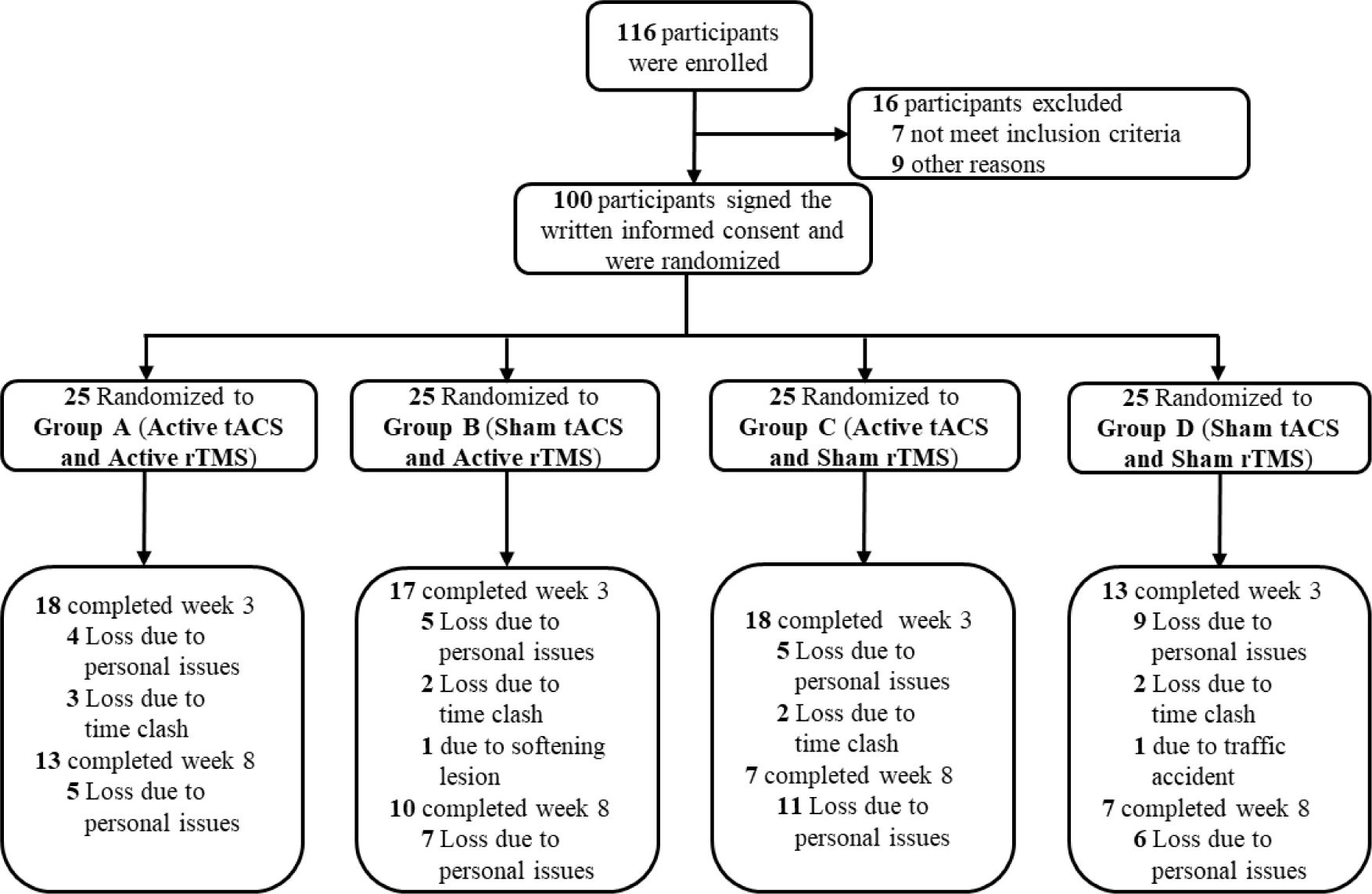
Flow diagram of participant selection and intervention

**Table 1.**
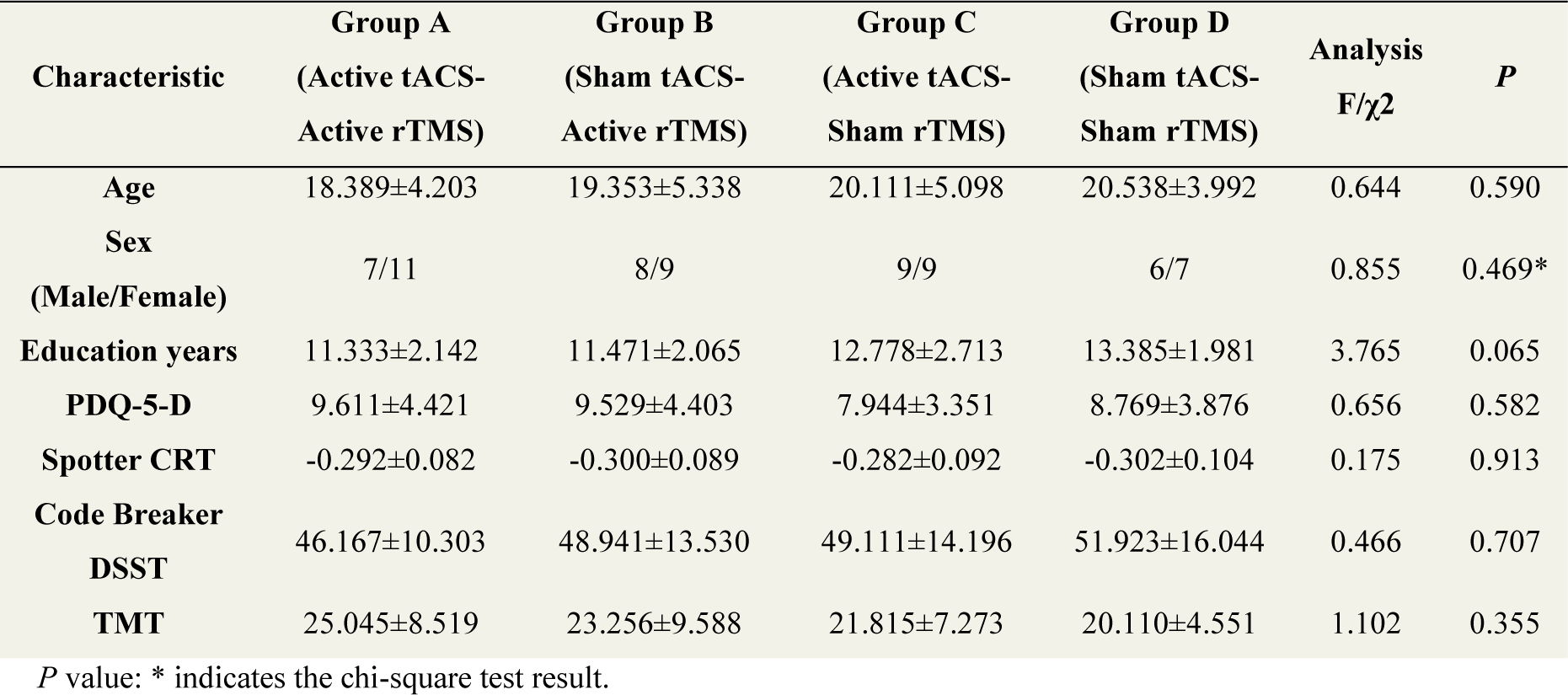
Demographic and clinical characteristics of the participants at baseline

### 3.2 Cognitive Function

A factorial design ANOVA was used to compare differences among the four groups pre- and post-intervention (Week 0 and Week 3). The analysis revealed a significant interaction effect between tACS and rTMS interventions on TMT and DSST metrics (*P* < 0.00001). As shown in Figure 4, after 3 weeks of continuous intervention, Group A demonstrated more significant improvements in Spotter CRT, TMT, and DSST compared to the other treatment groups. However, during the follow-up assessment 5 weeks after the intervention ended (Week 8), sustained cognitive improvements were only observed in the Spotter CRT metric (*P* < 0.01). These findings suggest that the combined MNS therapy has a synergistic effect on cognitive enhancement in BD patients.

**Figure 4.**
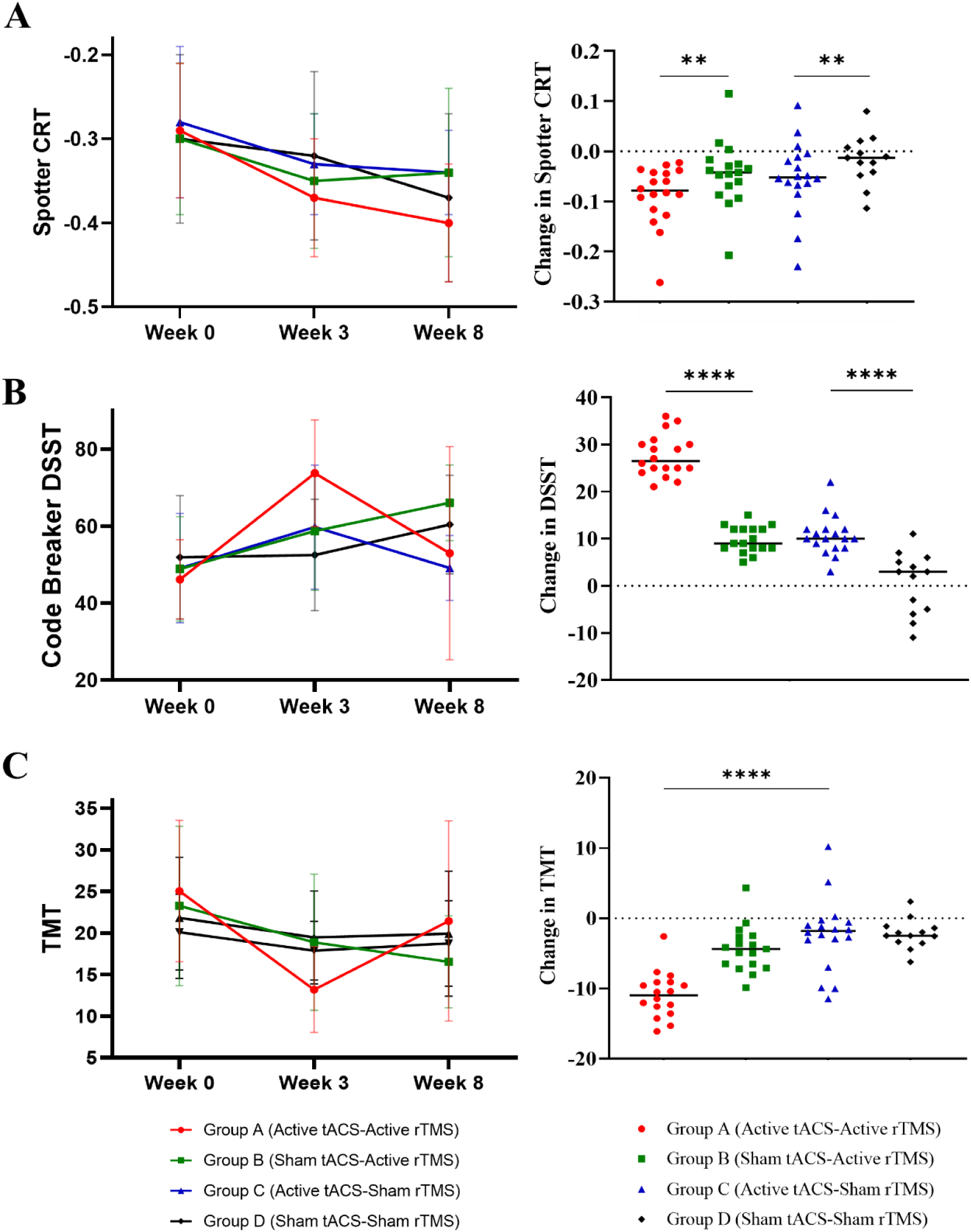
Clinical intervention results. The left panels show the scale scores of each intervention group at the three time points: W0, W3, and W8. The right panels display the changes in scores between W0 and W3. A.Results for the Spotter CRT. B.Results for the Codebreaker DSST. C.Results for the TMT. Each data point represents participants with recorded scores at that time point. W0: week 0, W3: week 3, W8: week8; \*\**P* < 0.01, \*\*\*\**P* < 0.0001.

### 3.3 fALFF Changes after MNS

After three-week intervention, we analyzed the resting-state functional indicators. Only the fALFF metrics in Group A passed the multiple comparison tests. As depicted in Figure 5, the left Frontal_Inf_Oper_L (inferior frontal gyrus, opercular part) showed significant activation following the combined tACS-rTMS intervention. This region is located near the targeted stimulation site, indicating a localized effect of the intervention.

**Figure 5.**
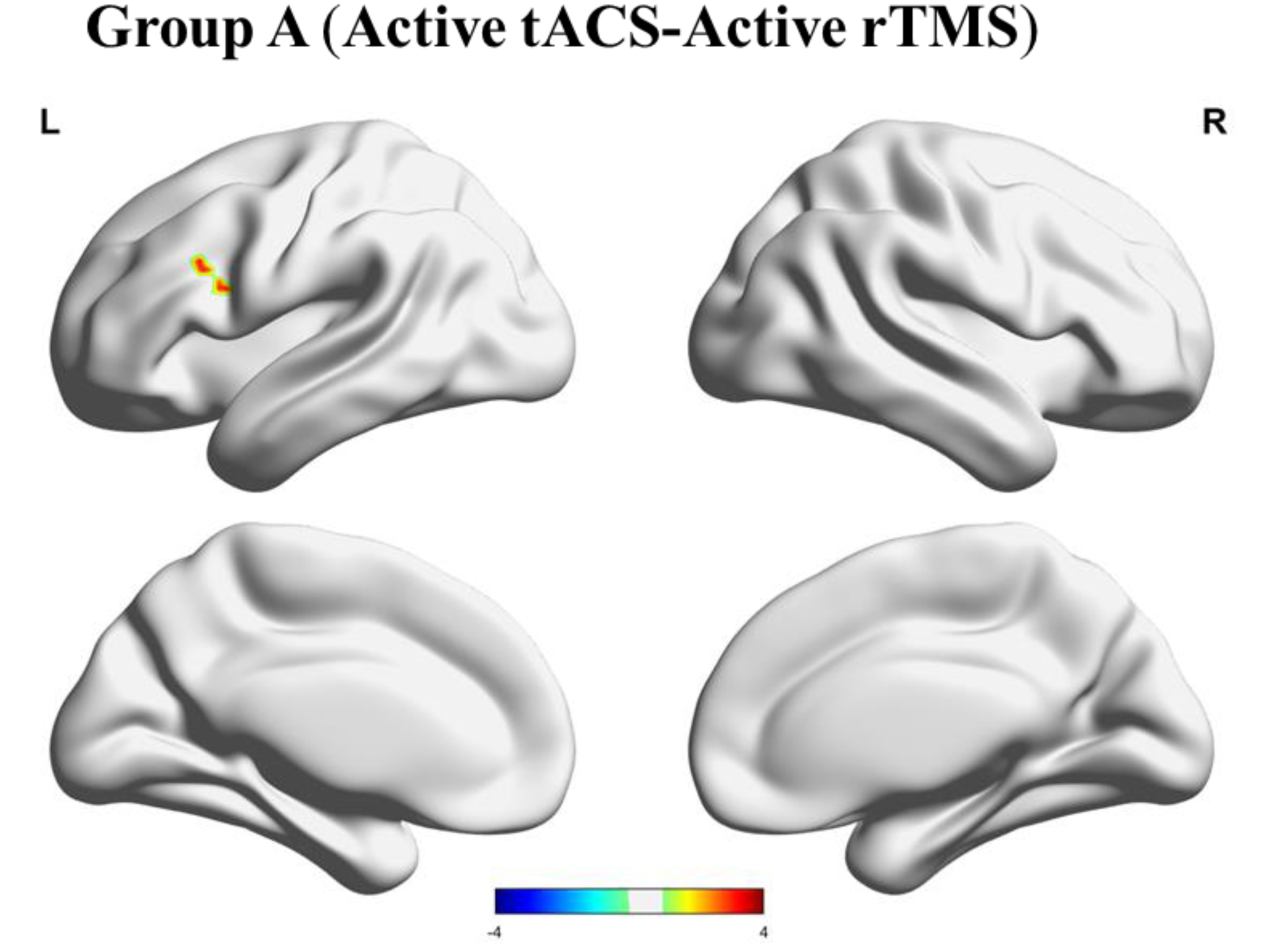
The spatial distribution of significant clusters in the brain. Red in the color bar indicates a significant increase in fALFF values at post-treatment compared to baseline (voxel-level threshold *p* < 0.01, cluster-level threshold *p* < 0.05, GRF corrected).

### 3.4 Adverse Effects

Adverse events were primarily observed in the immediate vicinity of and following the active tACS and rTMS interventions. During the tACS treatment, most patients reported experiencing a tingling sensation on the scalp, which was particularly noticeable during the first and last 10 seconds of the stimulation session. These symptoms were generally mild. Additionally, 12 patients experienced visual flickering. In the rTMS treatment phase, 8 patients reported transient dizziness following the session, which lasted approximately 5 seconds and did not affect their visual perception. These events were closely monitored and managed appropriately, with symptoms spontaneously resolving without further intervention.

## 4. Discussion

Through the implementation of a clinical randomized controlled trial, our novel Multimodal Neurostimulation (MNS) strategy has demonstrated significant clinical efficacy. The results indicate that the innovative MNS protocol is both effective and feasible for broader clinical application. The strategy combines electrical and magnetic stimulation while utilizing DTI for individualized target localization, presents a comprehensive approach that enhances therapeutic outcomes compared to traditional monomodal interventions.

A pivotal aspect of this study is the use of individualized DTI for precise target localization in BD patients. To our knowledge, this is the first study that employing DTI to identify intervention targets, specifically focusing on the densest white matter fiber tracts between the DLPFC and the dACC in BD patients. Physiologically, targeting these regions with high white matter fiber density could indicate stronger neural connections^47,48^, thereby offering a novel and potentially more effective method for target localization. This approach is similar to recent studies that use functional connectivity to determine individualized targets^29,30^, underscoring the importance of personalized target selection based on neural connectivity patterns to optimize therapeutic outcomes^49,50^.

Moreover, this study introduced individualized tACS with parameter optimization to maximize the electric field intensity at the target site. By creating personalized electric field simulations, tACS was fine-tuned to enhance the strength and focality of stimulation in the identified target area, potentially leading to better therapeutic outcomes. In addition, the use of robot-assisted rTMS ensured highly accurate and consistent localization of DTI-defined regions. This novel approach mitigates the inherent variability of manual targeting, ensuring that rTMS is precisely delivered to the intended neural circuits, thereby improving the reliability and efficacy of the treatment^51,52^.

The combined use of electrical and magnetic stimulation in a multimodal approach was employed to enhance intervention efficacy. The synergistic effects of electromagnetic co-stimulation have garnered increasing attention, with imaging results showing its impact on brain networks. Notably, even single-modal stimulation targeting DTI-identified sites resulted in cognitive improvements in patients, but the combined electromagnetic stimulation yielded superior outcomes. This observation supports the superiority of multimodal treatments, highlighting their significant clinical implications. However, several challenges remain, including determining the optimal duration and spatial configuration of electrical and magnetic interventions and assessing their effectiveness in treating other psychiatric conditions like treatment-resistant depression, warranting further exploration.

Despite its promising results, this study has certain limitations. Firstly, it was a single-center randomized double-blind study; future research should consider multicenter trials to validate these findings. Additionally, due to challenges in clinical data collection, post-intervention imaging data were not collected, limiting the analysis of long-term cumulative effects on brain networks. Consequently, future studies should incorporate longitudinal imaging to evaluate the sustained impact of MNS. Furthermore, while our approach has shown efficacy in targeting DLPFC and dACC, there is a need to explore its applicability across different neural circuits and conditions. Recent studies suggest that different psychiatric disorders may require tailored approaches based on their unique neural circuitry. Therefore, expanding the scope of individualized DTI-guided interventions across various disorders could provide a deeper understanding of the mechanisms underlying neuromodulation and its broader clinical applications.

## 5. Conclusion

This study proposes a novel DTI-guided MNS protocol that effectively improves cognitive function in BD patients. Our findings highlight the clinical relevance and applicability of this innovative MNS strategy, paving the way for new avenues in enhancing cognitive outcomes in psychiatric disorders.

## Conflict of Interest

All authors declare no competing interests.

## Data availability

Data involved in this study are available upon reasonable request.

## Acknowledgments

We would like to extend our heartfelt gratitude to all the patients who participated in this clinical study and their families for their invaluable support.

## Source of funding

Research supported by the National Key Research and Development Program of China (2023YFC2506200), the Research Project of Jinan Microecological Biomedicine Shandong Laboratory (No. JNL-2023001B), the Zhejiang Provincial Key Research and Development Program (2021C03107), the Leading Talent of Scientific and Technological Innovation—“Ten Thousand Talents Program” of Zhejiang Province (No. 2021R52016), the Innovation team for precision diagnosis and treatment of major brain diseases (No. 2020R01001), Chinese Medical Education Association (2022KTZ004), the National Natural Science Foundation of China (82201675) and the Fundamental Research Funds for the Central Universities (226-2022-00193, 226-2022-00002, 2023ZFJH01-01, 2024ZFJH01-01).

## Notes

### Competing Interest Statement

The authors have declared no competing interest.

### Clinical Trial

NCT05964777

### Author Declarations

The Clinical Research Ethics Committee of the First Affiliated Hospital, Zhejiang University School of Medicine gave ethical approval for this work (Approval number: IIT20230058C-R1).

